# Weak association of coinfection by SARS-CoV-2 and other respiratory viruses with severe cases and death

**DOI:** 10.1101/2020.07.22.20159400

**Authors:** L Fernandes-Matano, IE Monroy-Muñoz, LA Uribe-Noguez, MA Hernández-Cueto, B Sarquiz-Martínez, HD Pardavé-Alejandre, A Santos Coy-Arechavaleta, JE Alvarado-Yaah, T Rojas-Mendoza, CE Santacruz-Tinoco, C Grajales-Muñiz, VH Borja-Aburto, JE Muñoz-Medina

## Abstract

**Background:** SARS-CoV-2 is a novel coronavirus described for the first time in China in December 2019. This virus can cause a disease that ranges in spectrum from asymptomatic to severe respiratory disease with multiorgan failure, and the most severe cases are associated with some comorbidities and patient age. However, there are patients who do not have those risk factors who still develop serious disease.

**Methods:** In this study, we identified the presence of other respiratory viruses in positive cases of COVID-19 in Mexico to determine if any coinfections were correlated with more severe manifestations of COVID-19. We analysed 103 confirmed cases of COVID-19 using RT-qPCR for the detection of 16 other respiratory viruses.

**Results:** Of the cases analysed, 14 (13.6%) were cases of coinfection, and 92% of them never required hospitalization, even when comorbidities and advanced age were involved. There weren’t significant differences between the presence of comorbidities and the mean ages of the groups

**Conclusions:** These results suggest that coinfection is not related to more severe COVID-19 and that, depending on the virus involved, it could even lead to a better prognosis. We believe that our findings may lay the groundwork for new studies aimed at determining the biological mechanism by which this phenomenon occurs and for proposing corresponding strategies to limit the progression to severe cases of COVID-19.

**CLINICAL TRIAL REGISTRATION:** Not apply

## 1 INTRODUCTION

Coronaviruses (CoVs) circulate mainly in birds and mammals but can sometimes evolve, cross the barrier between species, and infect humans [1]. Until a few months ago, six different species of CoV were known to affect humans (human (H)CoV-229E, HCoV-NL63, HCoV-OC43, HCoV-HKU1, severe acute respiratory syndrome (SARS)-CoV, and Middle East Respiratory Syndrome CoV) [2]. However, in December 2019, cases of pneumonia without a known cause began to appear, epidemiologically linked to a fish and animal market in Wuhan, Hubei Province, China. Later, through the genetic sequencing of the isolates obtained from the patients, the aetiological agent was identified as a novel CoV species genetically close to CoVs of bats, which was named SARS-CoV-2, and the disease it causes was named coronavirus disease 2019 (COVID-19) [3-5].

On March 11, 2020, the WHO declared the outbreak of COVID-19 a pandemic, and by April 6, 2020, more than 1 million confirmed cases had been reported. The main clinical symptoms of the confirmed cases are fever, cough, shortness of breath, fatigue, and gastrointestinal symptoms [6].

This new disease can vary from asymptomatic to severe respiratory disease with multiorgan failure, and advanced age and some comorbidities, such as diabetes and hypertension, seem to be associated with more severe cases and death [6-8]. Nevertheless, there are cases of deaths from SARS-CoV-2 in various parts of the world that are not explained by the presence of these factors, so it is not yet clear what other factors could contribute to the development of more severe symptoms and the need for hospitalization. Some researchers propose, for example, that one of the causes of severe cases with no apparent explanation could be genetic predispositions linked to the gene encoding the cell-surface protein angiotensin-converting enzyme 2 (ACE2), used by SARS-CoV-2 to enter airway cells [9]. Our group was interested in investigating other variables that could explain such cases.

Because the pandemic caused by SARS-CoV-2 began during the influenza season, which is also the season with the highest incidence of various other respiratory viruses, we decided to evaluate whether the severity of confirmed cases of COVID-19 could be related to cases of coinfection.

## 2 MATERIALS AND METHODS

### 2.1 Study design

To evaluate whether coinfection with other respiratory viruses is associated with greater severity of COVID-19 cases in Mexico, we analysed samples of pharyngeal exudate received by the Central Laboratory of Epidemiology (CLE) that tested positive for SARS-CoV-2. As of March 30, 2020, CLE had received 1915 samples of suspected cases of COVID-19, of which 277 were positive. Of these, 103 had sufficient biological material to carry out the identification by RT-qPCR of another 16 respiratory viruses (human respiratory syncytial virus (HRSV), human parainfluenza viruses 1-4 (HPIV1-4), influenza virus A (Inf A), influenza B virus (Inf B), human mastadenovirus (HMdV), rhinovirus (RV), enterovirus (EV), human metapneumovirus (HMpV), primate bocaparvovirus (PBpV), HCoV-229E, HCoV-OC43, HCoV-NL63, and HCoV-HKU1).

Other factors that could contribute to the increased severity of the cases analysed were evaluated, such as age, sex, the presence of comorbidities (diabetes, hypertension, chronic kidney disease, chronic liver disease, chronic obstructive pulmonary disease (COPD), asthma, obesity, and haemolytic anaemia), and viral load at the onset of symptoms.

All 103 samples analysed had a previous positive result for SARS-CoV-2 by RT-qPCR and met the following case definition: a person of any age who in the last 7 days presented with at least two of cough, fever, and headache, accompanied by at least one of dyspnoea (severity data), arthralgias, myalgias, odynophagia/pharyngeal burning, rhinorrhoea, conjunctivitis, and chest pain. The other inclusion criterion was that the data on the patient’s initial clinical condition at the time of sample collection and the final outcome of the disease were available.

### 2.2 Extraction of nucleic acids

Total nucleic acids were obtained from 200-μL samples of pharyngeal exudate, taken with a Dacron swab (Copan Diagnostics, Corona, California, Catalogue: 159C), stored in viral transport medium (BD™ Universal Viral Transport System, East Rutherford, New Jersey, USA Catalogue: 220220) at −80 °C until use. For the extraction of total nucleic acids, the automated system MagNA Pure LC 2.0 (Roche Diagnostics, Mannheim, Germany) and the MagNA Pure LC Kit (Roche Diagnostics, cat. No. 03038505001) were used according to the manufacturer’s instructions.

### 2.3 Identification of SARS-CoV-2 and viral coinfections by RT-qPCR and qPCR

The viruses were evaluated according to the Guidelines for Laboratory Surveillance of Influenza and Other Respiratory Viruses from the Institute of Epidemiological Diagnosis and Reference (*Instituto de Diagnóstico y Referencia Epidemiológicos*) [10] and following the method of Corman et al. [11]. The SuperScript™ III Platinum® One-Step RT-qPCR System Kit (Invitrogen, Carlsbad, California, USA, Catalogue: 12574035) was used for the amplification of the viral genetic material in a 7500 Fast Real-Time PCR System (Applied Biosystems®, Foster City, California, USA). The viruses were evaluated in uniplex reactions with the following reaction mixture: 12.5 μl of 2x reaction mixture, 0.5 μl of each primer and probe, 0.5 μl of enzyme, 5.5 μl of RNase-free water, and 5 μl of total nucleic acid. The following thermocycling conditions were used: one cycle of 50 °C for 15 min and 95 °C for 2 min, followed by 45 cycles of 95 °C for 15 seconds and 60 °C for 30 seconds (SARS-CoV-2) or 55 °C for 1 min (other viruses). The sequences of the primers and probes, as well as the working concentration of each, are given in Table 1.

**Table 1.**
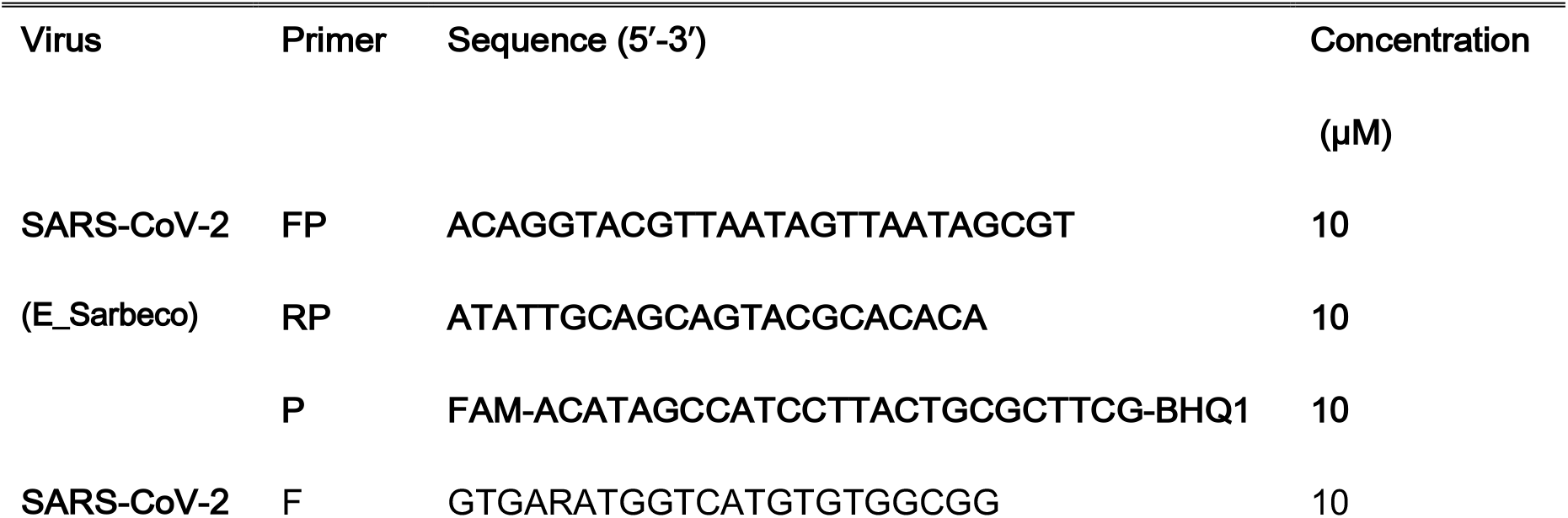
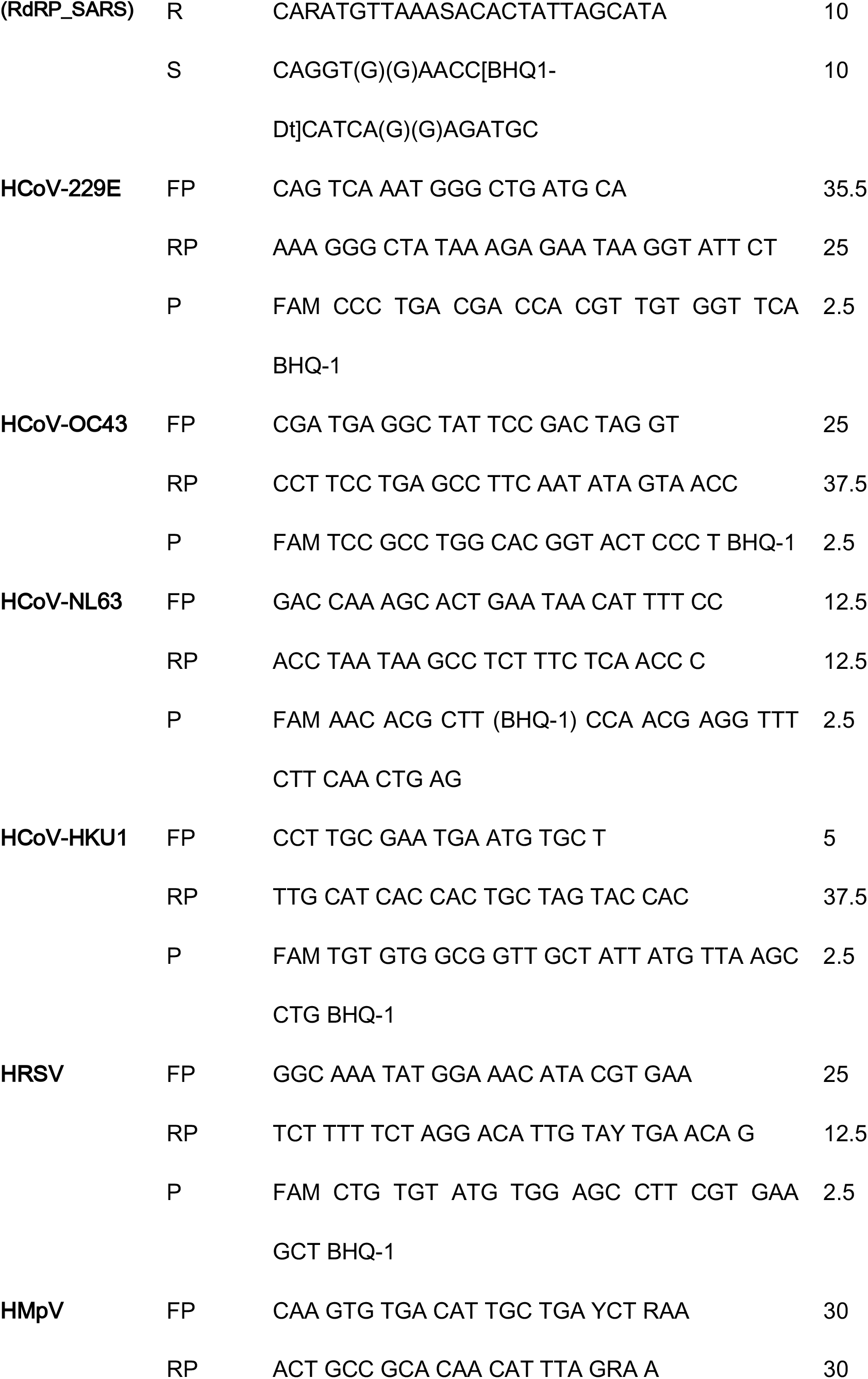
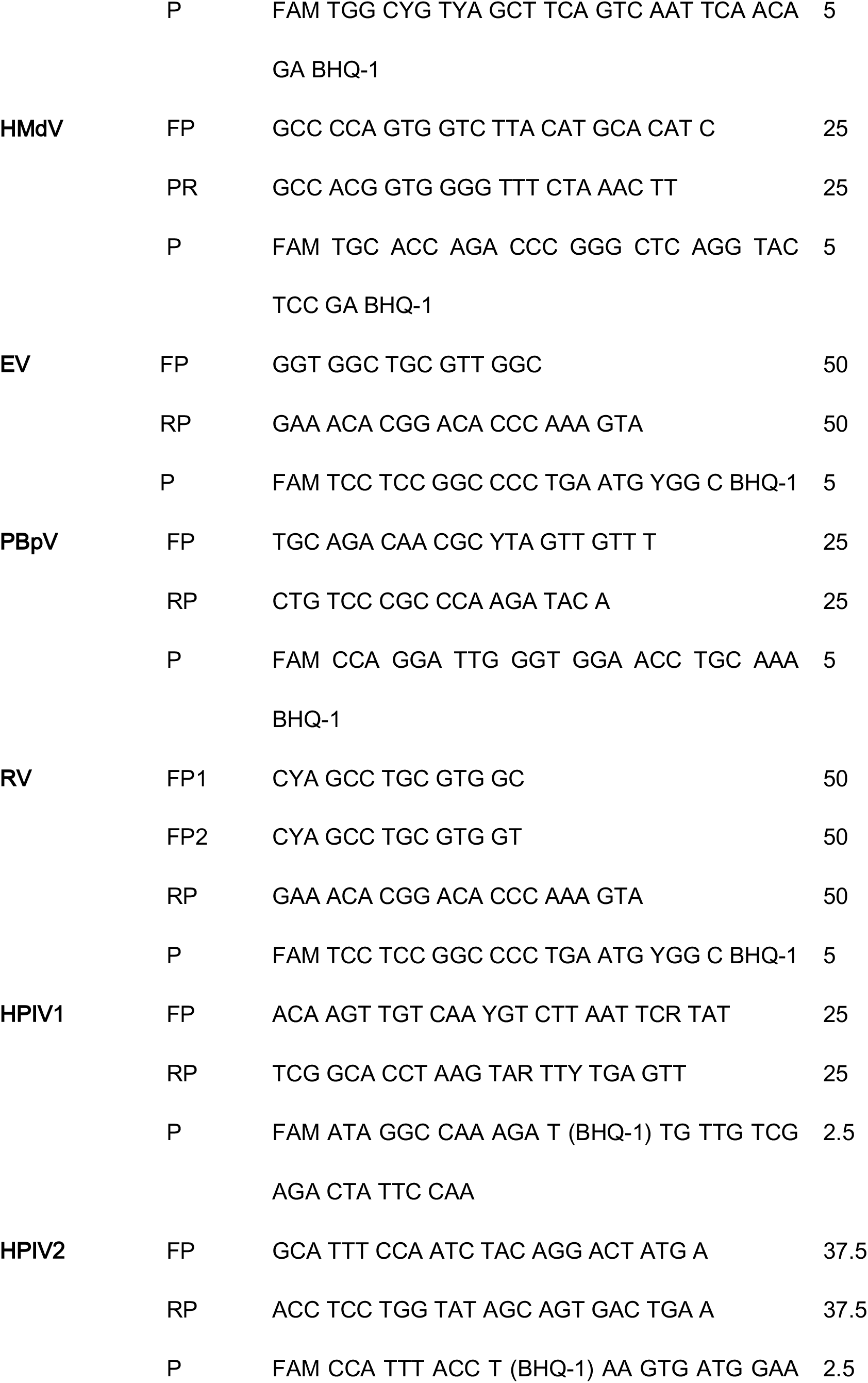
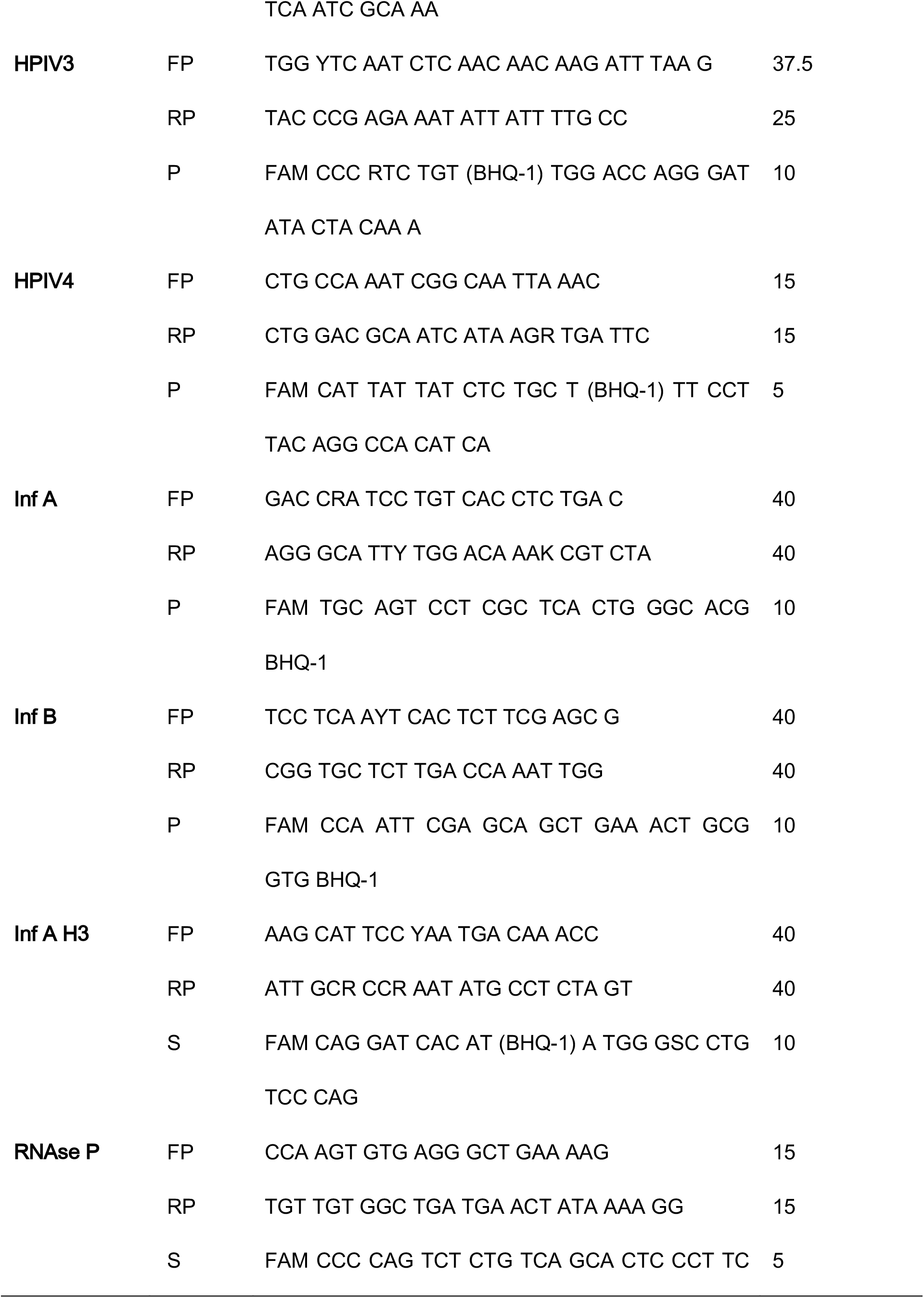
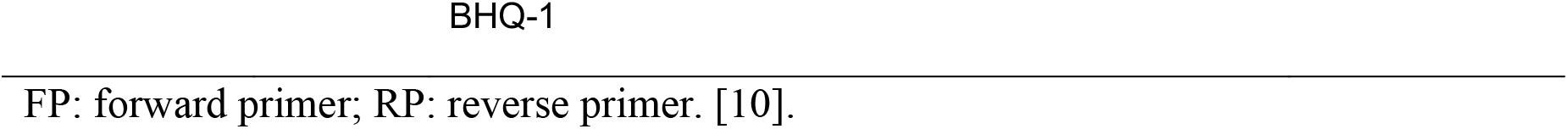
Sequences and working concentrations of the primers used for the detection of SARS-CoV-2 and 16 other respiratory viruses.

### 2.4 Reaction controls and interpretation

Lyophilised RNA and DNA (AmpliRun®, Vircell, Granada, Spain) were used as positive controls for all viruses tested. All samples showing amplification of any of the viral markers (cycle threshold <37) and the RNase P control were considered positive, and all samples without amplification of the viral markers but with amplification of the RNase P control were considered negative.

### 2.5 Quantification of the viral load of SARS-CoV-2

For the determination of the absolute viral load of SARS-CoV-2, a standard ten-point curve of the SARBECO_E region (Table 1) was generated using 1:10 dilutions from a stock of known concentration (1×10^10^ to 1×10^1^). For reverse transcription and amplification of the target region, the SuperScript™ III Platinum® One-Step RT-qPCR System Kit (Invitrogen, cat. No. 12574035) was used in a 7500 Fast Real-Time PCR System (Applied Biosystems®, Foster City, California, USA). The dynamic quantification range was from 10^10^ to 10^4^. The R^2^ value of the generated curve was 0.997.

### 2.6 Statistical analysis

Descriptive statistics are used to report frequencies, and the means are given with their 95% confidence intervals. The χ^2^ test of homogeneity and independence and Fisher’s exact test were used to compare categorical variables (P<0.05 was considered significant). Analysis of variance, Student’s t test, or the Mann-Whitney U test was used compare quantitative variables, as appropriate. The analyses were performed in IBM SPSS Statistics 24.0, and the graphs were generated with Microsoft® Excel® 2010.

## 3 RESULTS

### 3.1 Demographic analysis

Of the 103 cases of COVID-19 analysed, 74 (71.8%) were of outpatients who never needed hospitalization, 12 (11.7%) were of patients who recovered but who at some point had more severe symptoms and needed hospitalization in the course of the disease, and 17 (16.5%) were of patients who needed hospitalization but died. Sixty-two were men (60.2%), and 41 were women (39.8%). The mean age was 44.4 years, ranging from 6 to 85. The population was divided into four age groups according to the Mexican Institute of Social Security’s health handbooks.

Following this classification, the cases were distributed as follows: one patient was aged 0 to 9 years (0.9%), one aged 10 to 19 years (0.9%), 83 aged 20 to 59 years (80.6%), and 18 aged 60 years or older (17.5%). The samples analysed were from 18 Mexican states; 13 were from the northern region (12.6%), 80 the central region (77.7%), and 10 the southern region (9.7%) (Table 2).

**Table 2.** Demographic data of the cases included in the study.

|  | Total | Outpatients | Hospitalized | Deaths |
| --- | --- | --- | --- | --- |
|  | N=103 | N=74 | N =12 | N =17 |
|  | n (%) | n (%) | n (%) | n (%) |
| **Sex** |  |  |  |  |
| Male | 62 (60.2) | 39 (52.7) | **9 (75.0)\*** | **14 (82.4)\*** |
| Female | 41 (39.8) | 35 (47.3) | 3 (25.0) | 3 (17.6) |
| **Age group** |  |  |  |  |
| 0-9 years | 1 (0.9) | 0 (0.0) | 1 (8.3) | 0 (0.0) |
| 10-19 years | 1 (0.9) | 1 (1.3) | 0 (0.0) | 0 (0.0) |
| 20-59 years | 83 (80.6) | 64 (86.5) | 8 (66.7) | 11 (64.7) |
| ≥60 years | 18 (17.5) | 9 (12.2) | 3 (25.0) | 6 (35.3) |
| Mean age (years) | 44.4 | 41.6 | 46.5 | **55.2\*** |
| **Region** |  |  |  |  |
| North | 13 (12.6) | 10 (13.5) | 0 (0.0) | 3 (17.6) |
| Central | 80 (77.7) | 57 (77.0) | 11 (91.7) | 12 (70.6) |
| South | 10 (9.7) | 7 (9.5) | 1 (8.3) | 2 (11.8) |

N: total analysed samples; n: identified samples. \*P<0.05.

Table 2 also shows that the percentages of hospitalizations and deaths were significantly higher in male patients than in female patients (both P<0.05) and that older age was also associated with a higher probability of death due to COVID-19, with the mean age in the group of outpatients being 41.6 years versus 55.2 years in the group of deaths (P<0.05).

### 3.2 Coinfections

In the study, 14 cases of coinfections were identified, equivalent to 13.6% of the total samples analysed. The most detected viruses were HCoV-229E and HRSV, adding up to 57.1% of the total co-infections. Besides HCoV 229E, another coronavirus was detected, HCoV-OC43, which belongs to the same genus as SARS-CoV-2. The rest of the cases involved HRV, Inf A, HPIV1, HPIV2, and HPIV4 (Fig. 1).

**Fig 1.**
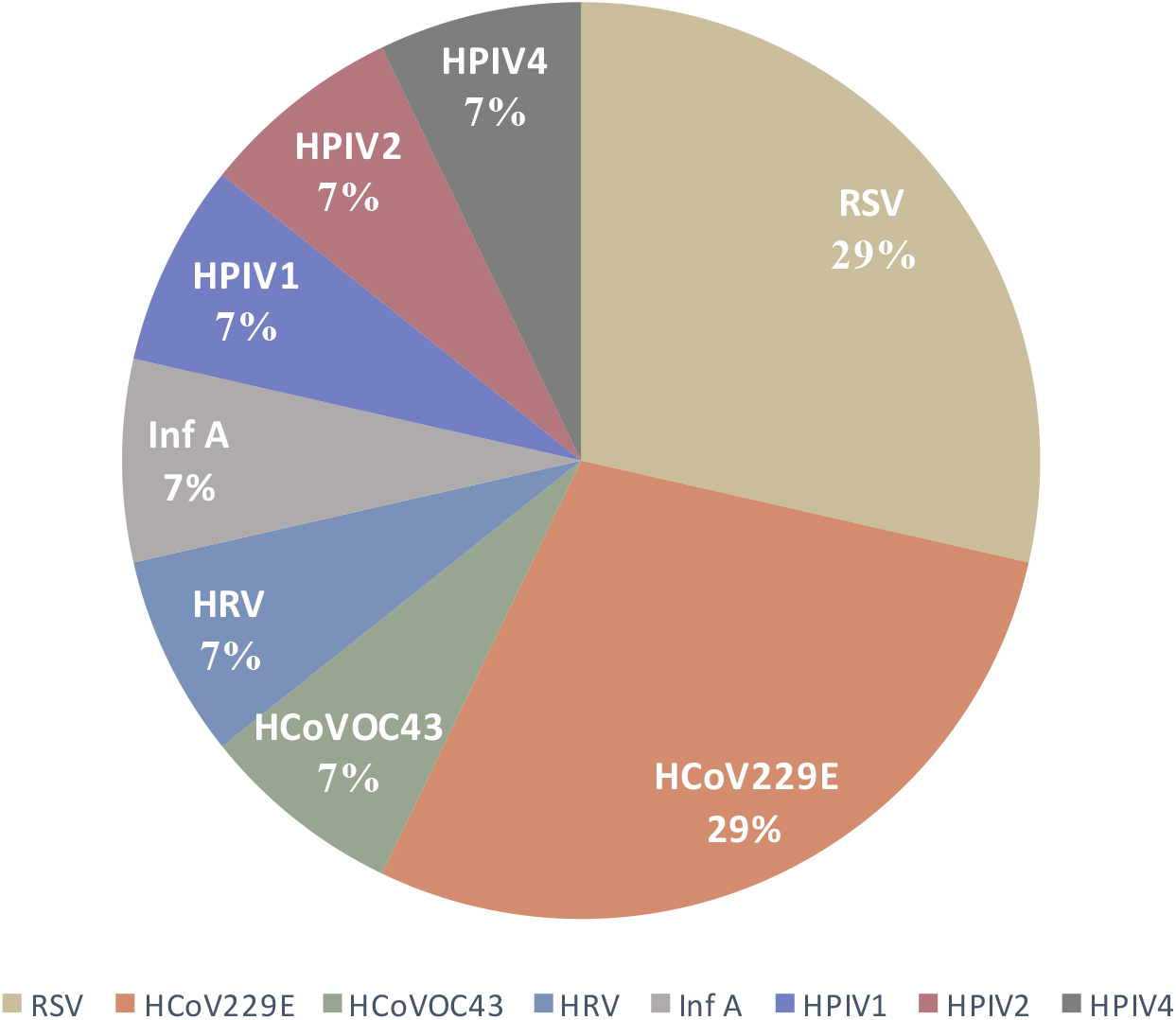
Viruses involved in coinfections with SARS-CoV-2.

In general, the coinfected patients presented a milder picture of respiratory infection, 13 (92.9%) of them corresponding to outpatient cases. Only the case of coinfection involving Inf A required hospitalization and resulted in the death of the patient.

The symptoms presented by the coinfected patients are shown in Table 3. No significant differences were found between the initial symptoms of these patients and the symptoms of those infected only with SARS-CoV-2, with the exception of diarrhoea. There were also no differences in the number of symptoms between the two groups. It is worth mentioning that the most severe symptoms, such as dyspnoea and the few cases of cyanosis, occurred almost exclusively in patients infected only with SARS-CoV-2.

**Table 3.** Comparison of the symptoms presented between patients with coinfection and single infection by SARS-CoV-2.

| Symptom | Total<br>N=103<br>n (%) | Coinfection<br>N=14<br>n (%) | Single infection by<br>SARS-CoV-2<br>N=89<br>n (%) |
| --- | --- | --- | --- |
| Cough | 87 (84.5) | 11 (78.6) | 76 (85.4) |
| Fever | 84 (81.6) | 12 (85.7) | 72 (80.9) |
| Headache | 84 (81.6) | 12 (85.7) | 72 (80.9) |
| Myalgias | 72 (69.9) | 11 (78.6) | 61 (68.5) |
| Arthralgias | 65 (63.1) | 9 (64.3) | 56 (62.9) |
| Odynophagia | 57 (55.3) | 8 (57.1) | 49 (55.1) |
| Chills | 55 (53.4) | 10 (71.4) | 45 (50.6) |
| Rhinorrhoea | 51 (49.5) | 10 (71.4) | 41 (46.1) |
| Chest pain | 40 (38.8) | 5 (35.7) | 35 (39.3) |
| Dyspnoea | 30 (29.1) | 2 (14.3) | 28 (31.5) |
| Diarrhoea | 23 (22.3) | **6 (42.9)\*** | 17 (19.1) |
| Abdominal pain | 22 (21.4) | 3 (21.4) | 19 (21.3) |
| Conjunctivitis | 13 (12.6) | 3 (21.4) | 10 (11.2) |
| Adynamia | 11 (10.7) | 2 (14.3) | 9 (10.1) |
| Coryza | 6 (5.8) | 2 (14.3) | 4 (4.5) |
| Cyanosis | 5 (4.9) | 0 (0.0) | 5 (5.6) |
| Polypnea | 5 (4.9) | 0 (0.0) | 5 (5.6) |
| Dysphonia | 0 (0.0) | 0 (0.0) | 0 (0.0) |
| Nasal congestion | 0 (0.0) | 0 (0.0) | 0 (0.0) |
| Low back pain | 0 (0.0) | 0 (0.0) | 0 (0.0) |

N: total analysed samples; n: identified samples. \* P<0.05.

To verify whether the coinfected patients presented milder cases of the disease due to a better general health status, we compared the single-infected and coinfected groups on age, several comorbidities, and immune status. The comorbidities evaluated were diabetes, hypertension, chronic kidney disease, chronic liver disease, COPD, asthma, obesity, and haemolytic anaemia. The statistical analyses did not reveal significant differences in the frequency of any of these comorbidities between the two groups (P>0.05) (Table 4).

**Table 4.** Comparison of comorbidities and age between SARS-CoV-2 coinfection and single-infection groups.

|  | **Total** | **Coinfection** | **Single infection by** |
| --- | --- | --- | --- |
|  | **N=103** | **N=14** | **SARS-CoV-2** |
|  | **n (%)** | **n (%)** | **N=89** |
|  |  |  | **n (%)** |
| **Comorbidities** |  |  |  |
| Obesity | 21 (20.4) | 4 (28.6) | 17 (19.1) |
| Hypertension | 19 (18.4) | 4 (28.6) | 15 (16.9) |
| Diabetes | 16 (15.5) | 3 (21.4) | 13 (14.6) |
| Asthma | 5 (4.9) | 0 (0.0) | 5 (5.6) |
| COPD | 5 (4.9) | 0 (0.0) | 5 (5.6) |
| Chronic kidney disease | 2 (1.9) | 0 (0.0) | 2 (2.2) |
| Chronic liver disease | 0 (0.0) | 0 (0.0) | 0 (0.0) |
| Haemolytic anaemia | 0 (0.0) | 0 (0.0) | 0 (0.0) |
| **Immunosuppression** | 3 (2.9) | 1 (7.1) | 2 (2.2) |
| **Mean age** | 44.4 ($\pm 3.0$) | 49.4($\pm 7.3$) | 43.6($\pm 3.2$) |

N: total analysed samples; n: identified samples.

Next, to verify whether coinfection with other respiratory viruses positively or negatively affected the viral load of SARS-CoV-2, absolute quantification was performed by RT-qPCR in all the samples included in the study. Although the P-value was not significant (P>0.05), the mean viral load was lower in the cases of coinfection (676,220 ± 749,715 vs 2,353,580 ± 1,283,262) (Fig. 2). Both the viral load, as well as the age and comorbidities of each coinfected patient can be seen in Table 5.

**Fig 2.**
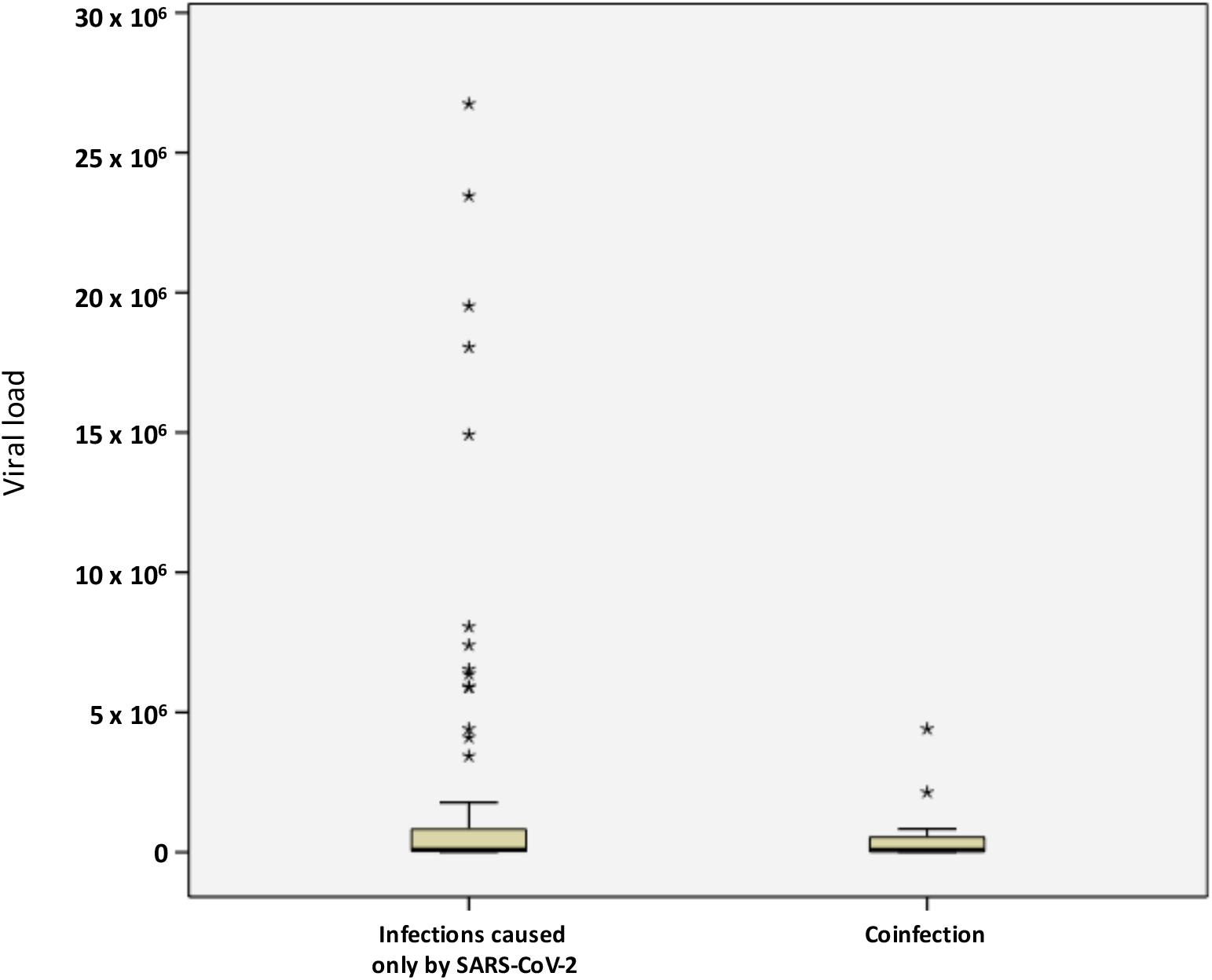
Comparison between viral load in cases of coinfection and infections caused only by SARS-CoV-2. Only the samples in which the viral load could be determined were plotted (85 of 103) and one extreme point has been removed from each group, which differed by more than 25 million copies from the nearest point of each group.

**Table 5.** Summary of the characteristics of the coinfection cases.

| CASE | Virus | Viral load | Age | Comorbidities | Patient type |
| --- | --- | --- | --- | --- | --- |
| 6 | RSV | 4,404,336 | 60 | OB, HP | OUTPATIENT |
| 15 | RSV | 228,723 | 73 | - | OUTPATIENT |
| 21 | HCoV 229E | 11,197 | 33 | - | OUTPATIENT |
| 23 | HRV | <10000 | 50 | - | OUTPATIENT |
| 40 | HCoV 229E | 138,944 | 47 | - | OUTPATIENT |
| 42 | HCoV OC43 | 2,140,772 | 49 | - | OUTPATIENT |
| 45 | HCoV 229E | 11,956 | 53 | - | OUTPATIENT |
| 48 | HCoV 229E | 15,140 | 37 | - | OUTPATIENT |
| 56 | HPIV4 | 88,214,714 | 51 | - | OUTPATIENT |
| 59 | HPIV2 | 827,125 | 37 | DB, INMS, OB, HP | OUTPATIENT |
| 60 | RSV | 10,215 | 21 | - | OUTPATIENT |
| 77 | Inf A | 48,974 | 53 | DB, OB | DEATH |
| 100 | HPIV 1 | 244,226 | 58 | DB, OB, HP | OUTPATIENT |
| 101 | RSV | 33,042 | 69 | HP | OUTPATIENT |

## 4 DISCUSSION

In Mexico, as in many parts of the world, respiratory infections are the health conditions with the highest morbidity and mortality each year [12,13]. According to previous studies conducted by our research group, there is evidence that at least 16 respiratory viruses co-circulate in Mexico (RV, HRSV, HMpV, HMdV, EV, PBpV 1, HPIV 1, HPIV 3, HPIV 4, HCoV-229E, HCoV-OC43, HCoV-NL63, HCoV-HKU1, Inf A/H1N1pdm09, Inf A/H3N2, and Inf B) [14,15], aside from the recently introduced SARS-CoV-2. These viruses frequently cause cases of coinfections within the Mexican population, where the same patient can be infected with up to four different respiratory viruses at the same time [14].

Since shortly after the identification of this novel virus in Wuhan, China, demographic groups and risk factors have been reported that predispose to a more severe COVID-19 progression, but serious cases and even deaths that are not explained by these factors continue to appear. In this study, we verified the presence of 16 respiratory viruses in COVID-19-positive patients in Mexico to determine the existence of coinfections, as well as the relationship that coinfections may have with more severe manifestations of the disease.

After analysing 103 samples, 14 coinfections (13.6%) were identified. This percentage was slightly lower than that found by Navarro-Marí et al. [16] in Spain, who retrospectively looked for the presence of other respiratory viruses during the 2009 influenza pandemic and reported 15% coinfection in the more than 18,000 analysed samples [15]. Although their study addressed other important topics on the behaviour of some respiratory viruses, the clinical data associated with coinfections were not analysed [16].

In our study, contrary to what would be expected, the group of patients with co-infections presented evidently less severe manifestations of COVID-19, since 92.9% of these cases developed mild forms of the disease and the patients did not require hospitalization (outpatient cases). Additionally, comorbidities considered risk factors were equally prevalent in both groups, and the two groups had similar mean ages (Table 4), which shows that both groups presented the same predisposition to developing complications.

Of all the coinfections detected, only the case of coinfection with influenza A resulted in the death of the patient, which led us to investigate antecedents on whether the severity of the coinfection could depend on the specific type of virus that causes it or what other factors could explain our findings. In 2016, Pinky and Dobrovolny published one of the few existing studies on coinfections with different combinations of respiratory viruses. In their study, using a mathematical model, they demonstrated that viral interference can be explained mainly by competition for the resources of the host cell. Apparently, this interference also depends on which infection occurs first, and the authors predicted that viruses with higher growth rates can outperform viruses with lower growth rates, since the fastest-growing virus would consume more target cells at the beginning of infection [17].

When measuring the SARS-CoV-2 viral load in the cases analysed, we observed, although without statistical significance (P>0.05), that the mean viral load was lower in the coinfected patients (Fig. 2). This could suggest that there are viruses whose replication rates are higher than those of SARS-CoV-2, directly influencing its replication and resulting in a milder case of COVID-19. Even though we cannot affirm that the virus with a higher replication rate is the one that dictates the clinical manifestations of the coinfected patient, it seems not to be random that the seven viruses that were detected simultaneously in mild cases of COVID-19 (HCoV-229E, RSV, RV, HPIV1, HPIV2, HPIV4, and HCoV-OC43) cause generally self-limited respiratory infections that resolve in an average of 15 days and that, unlike these viruses, Inf A is associated with higher rates of complications and mortality than the other viruses and may have been the cause of death of the coinfected patient.

In any case, the interactions of two or more viruses when they simultaneously infect a host are more complex than they seem. In addition to competition for the resources of the host cell, other mechanisms are linked to the immune response and production of cytokines, such as interferons, that may also have some role in the viral interference phenomenon [18-20].

Therefore, larger and more targeted studies are required to corroborate the observations described in this work and to determine if coinfection with some viruses could lead to a better prognosis for patients diagnosed with COVID-19.

## 5 CONCLUSIONS

Despite the limitation of the small sample size, our results suggest that coinfection by some respiratory viruses and SARS-CoV-2 could mitigate the viral load of this novel virus and thus lead to a less severe clinical picture. We believe that our findings may lay the groundwork for new studies aimed at determining the biological mechanism by which this phenomenon occurs and for proposing corresponding strategies to limit the progression to severe cases of COVID-19.

## Data Availability

All relevant data are within the paper and the information is supported by supplementary files that can be consulted with the corresponding author.

## ACKNOWLEDGEMENTS

**We thank all the staff of the Molecular Biology and Sample Reception Departments at Central Laboratory of Epidemiology for their support**.

## FUNDING

**None**

## ROLE OF THE FUNDING SOURCE

**None**

## CONFLICT OF INTEREST

**The authors have no conflicts of interest to declare**

## ETHICS APPROVAL

**Human serum specimens were an excess of sample collected during routine passive surveillance activities of the Central Laboratory of Epidemiology**. **All specimens were delinked from any personal identifiers prior to commencement of the study**. **All the samples were used in an anonymous way**.

## INFORMED CONSENT

**Not apply**.

## DATA AVAILABILITY

**All relevant data are within the paper and the information is supported by supplementary files**.

## ABBREVIATIONS

**None**

## Notes

### Competing Interest Statement

The authors have declared no competing interest.

### Funding Statement

No external funding was received.

### Author Declarations

División de Laboratorios de Vigilancia e Investigación Epidemiológica.

